# Helping Breast Cancer Diagnosis on Mammographies using Convolutional Neural Networks

**DOI:** 10.1101/2024.07.31.24311257

**Authors:** Rocío García-Mojón, Fernando Martín-Rodríguez, Mónica Fernández-Barciela

## Abstract

In this paper a study about breast cancer detection is presented. Mammography images in DICOM format are processed using Convolutional Neural Networks (CNN’s) to get a pre-diagnosis. Of course, this preliminary result needs to be checked by a trained radiologist. CNN’s are trained and checked using a big database that is publicly available. Standard measurements for success are computed (accuracy, precision, recall) obtaining outstanding results better than other examples from the literature.

## 1. Introduction

### 1.1. Problem definition

According to the World Health Organization (WHO), breast cancer is the most frequent type of cancer worldwide, with more than 2.2 million cases in 2020 [1]. About 1 out of 12 women will suffer breast cancer during her life. In addition, it affects to all age ranges from puberty, although probability increases from 40 years old. About half cases of breast cancer correspond to women with no identifiable risk factor (excepting gender and age). Some circumstances to consider are obesity, or high alcohol consumption, family history, radiation exposure, tobacco consumption, post-menopause hormone therapy, and reproductive history.

Breast cancer is a malignant proliferation of the epithelial cells covering the mammary ducts that leads to tumor formation. There are two main types of breast cancer, which are infiltrating ductal carcinoma (approximately 80% of cases) and infiltrating lobular carcinoma (10-12% of cases), although there are others (which do not exceed 10% of the total). The most commonly used test for the diagnosis of this disease is breast radiography (mammography) because it is a minimally invasive method. From there, if necessary, other tests are performed. In the last case a biopsy is the test that allows a definite diagnosis.

Since 1980, in the most developed countries, mortality has been reduced by 40% due to the implementation of regular mammograms in risk groups. Therefore, early detection and treatment is very important to reduce the number of deaths. In addition, there are huge disparities in patient survival between high-income countries and middle and low-income ones, due to the cost of diagnosis, which requires the participation of radiologists with expertise in this field. These physicians look for small white spots (calcifications), large abnormal areas, and other suspicious areas in the images. In addition, the density of the breast also affects the identification of cancer, being more difficult in dense breasts. Furthermore, diagnostic tests of this type lead to a high incidence of false positives, causing unnecessary anxiety, additional tests and waiting in patients.

The described facts motivate researchers to design new tools for the diagnosis of breast cancer using screening mammograms. One of the existing possibilities is the automatic treatment of this type of medical images by performing a “pre-diagnosis” that can be confirmed or not by the specialist. This requires the use of automatic learning techniques (ML, Machine Learning). Currently, automatic image recognition is most frequently based on a mature technique known as Convolutional Neural Networks (CNN) [1]. This tool is a type of Deep Learning technique that, based on training data, extracts features that will be used to perform a classification. Compared to other older techniques, such as MLP (Multi-Layer Perceptron [2]) neural networks, SVM’s (Support Vector Machines [3]) or decision trees (such as Random Forest [4] or Bagged Tree: “Bootstrap Aggregated Tree” [5]). The advantage of CNN is that it does not require a manual selection of features or the design of procedures for their extraction. In classical techniques, features are selected by statistical measures of their relevance, called feature engineering techniques (techniques such as ANOVA or Pearson’s correlation coefficients [6]) or they are extracted by linear filters (technique preferably used in the case of images). The problem is that there is no systematic method for the design of these filters and researchers usually have acted based on experience. In a CNN, the first stages are convolutions (linear filters) where the coefficients are included in the training, i.e., the training designs the feature extraction. The last stages of a CNN (usually called “Fully Connected Layer”) are, basically, a multilayer perceptron (MLP).

### 1.2. Machine Learning

The proper treatment of data brings multiple benefits in all fields, since it helps decision making, reduces costs, improves market knowledge, etc. In addition, in the clinical field, the creation of tailored algorithms helps the early diagnosis of diseases, control of epidemics, prediction of hospital admissions, reduction of medical errors, etc. These advantages help researchers to face complex and challenging problems when managing these data.

Machine Learning is a branch of artificial intelligence that, from data and through a set of techniques, gives machines the ability to automatically learn a set of rules. This differs from classical programming, which consists in the execution of predetermined rules [7]. Machine Learning can help humans in routines such as speech recognition, image understanding, data analysis, weather forecasting, genomic analysis, e-commerce, etc. [8].

Deep Learning is known as the set of Machine Learning techniques, based on neural networks that are structured in different specialized levels (we are talking about more than two levels, since with only two we would have the classic MLP networks, now known as Shallow Learning systems). Normally, the first levels will extract certain characteristics or key parameters from the initial data and the last ones will make the relevant decisions. Examples of Deep Learning are convolutional neural networks (CNN), variants of these such as region-based convolutional networks (R-CNN [9]), networks designed for segmentation tasks such as U-NET [10] (an example of a semantic convolutional network) and, finally, networks designed to process sequences (such as audio signals or electrocardiograms) highlighting the LSTM (Long Short-Term Memory [11]).

During the last decade, deep learning has become one of the most valuable techniques of artificial intelligence and machine learning, showing a supreme performance in different areas, such as acoustics, image processing, and language processing. The main advantage of using Deep Learning applied to this particular field of medical imaging is that it complements human intelligence with the special capabilities of computers.

CNN’s are based on the working of biological neural networks. These receive the nervous impulse emitted by another neuron, process it and transmit a new impulse to the adjacent neurons with which they are connected. In the case of artificial neurons, these connections have numerical weights that adapt according to experience (training). Thus, the sum of the inputs, multiplied by their associated weights, determines a value that is processed in the activation function and sent as neuron output. The architecture of a CNN would be composed of different layers, in general, an input and an output layer, convolutional and activation layers and other intermediate layers that condense the image to improve feature extraction. A scheme can be seen in Figure 1.

**Figure 1.**
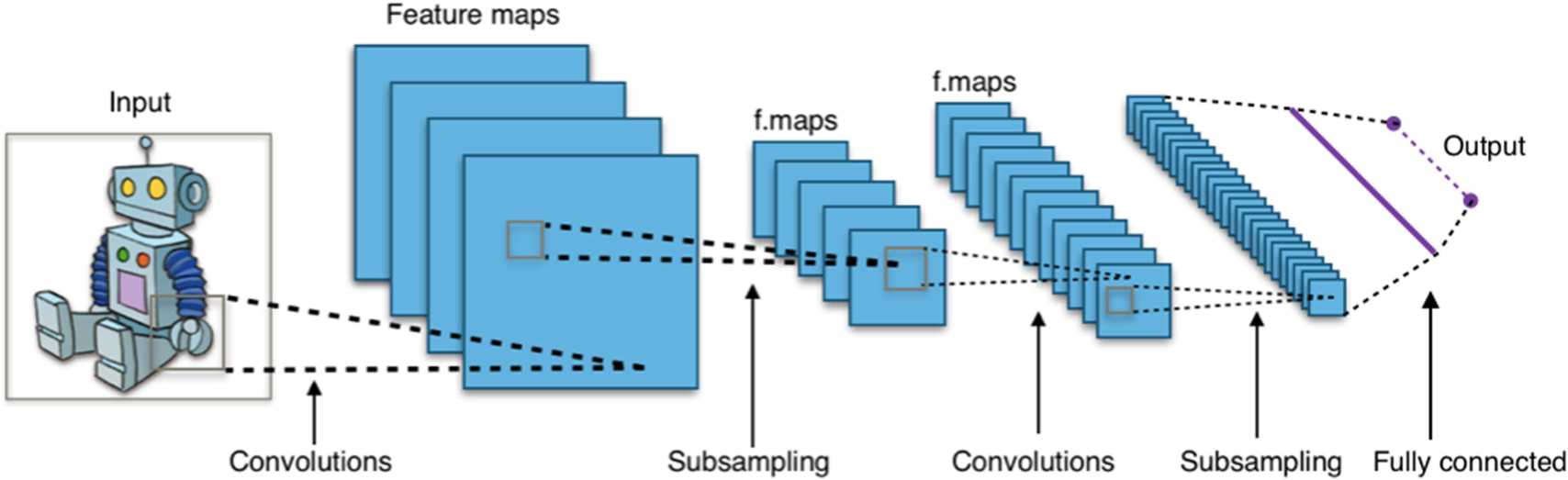
Typical CNN architecture [12]

### 1.3. Literature review

Some concrete and recent applications of CNNs in the medical field are, for example, diagnosis of Alzheimer’s disease and prediction of the progression of intellectual decay [13], lung cancer detection and classification [14, 15], detection of injuries on chest X-ray [16], identification and grouping of skin diseases [17], even the detection of certain diseases in crops [18]. There are also recent studies and CNN designs in the specific field to be addressed, Id. EST: breast cancer detection. Abdulla et al. obtained a 97.10% accuracy rate in their model, processing mammograms in order to eliminate the pectoral muscle and using SMOTE to introduce new samples for minority classes [19]. Tariq et al., propose a computer-aided diagnosis (CAD) system for breast cancer detection [20]. Alshehri et al., design a CNN that, after being trained with breast thermography images, obtains a rate of 92.32% [21]. Sakthivel et al. compare different methods of oversampling with a breast radiographies dataset and, after training the CNN, they find that the success rate increases between 19 and 34% [22].

In [19], [20] e [22] authors use the mini-MIA database consisting of 322 images so the results will be less significant than in this study (see information about the dataset in the next section). In [21], the authors use thermographs instead of mammograms; therefore, it is a different type of study.

It should also be noted that some results can achieve be very good measurements in terms of success rate (or accuracy) which is computed by dividing the number of correct recognitions by the total number of cases tested. However, this way of evaluating a system is not complete, especially in cases of unbalanced a priori probability between classes. This is: if before taking any mammography the probability of cancer is 10%, a system that produces always the result “absence of cancer” would be correct in 90% of cases. Therefore, more precise measures will be used in this study. This is the pair of values called: precision and recall [23]. Precision is the number of true positives versus the total number of positives obtained. This value indicates the trustworthiness of a positive result. While recall is the number of true positives against the total number of cancer cases (verified cases in the dataset). This last parameter is the most important in a diagnosis system, and if it is high it indicates that it is very unlikely to obtain a negative for a real cancer. These two values are grouped into a single number called F_1_-score which is the harmonic mean of both quantities (the inverse of the mean of the inverses). The harmonic mean is much more demanding than the arithmetic mean; a decrease in either of the two initial parameters will cause a strong drop in the F_1_-score.

In references [19] and [20], authors do not use these measurements. Reference [21] uses them for thermographs. In [22] F-score is computed but using another definition because recall is not computed in this reference, so it is difficult to compare results.

### 1.4. Class unbalance

Currently one of the main challenges and a key factor in the treatment of this data in the clinical field is the high unbalance between the classes. In this way, it is common to find an unbalanced dataset, this means: with a large number of negative cases and few positive ones. Therefore, a series of methods will have to be applied to balance both classes.

Traditionally, the methods used to attack this issue consisted of balancing the representation of the classes, either by multiplying the examples of the minority class (*oversampling*, creating new samples equal or similar to the original ones), or on the contrary, removing samples of the majority class (*undersampling*). Sometimes, both can be used at the same time; until reaching a balance in the number of samples per class. For example, SMOTE is a recent oversampling technique, based on generating new synthetic samples from the minority class samples [24].

With the aim of diagnosing breast cancer accurately and reducing the use of more invasive techniques, this work proposes a model for early detection of breast cancer through the application of Deep Learning. Specifically, the design of a CNN for the classification of mammograms according to whether they are healthy (without cancer) or sick (with cancer). This paper describes: first, the processing and preparation of the input data for its introduction into the automatic learning model; next, the experiments that were carried out to develop different CNN architectures, as well as the assessment and obtained results.

The global purpose is creating a computer application that can perform an automatic pre-classification, indicating to the radiologist which patients should have special attention and therefore helping in the diagnosis process.

## 2. Materials and Methods

In order to improve the accuracy and efficiency of the diagnosis carried out by radiologists, as well as the quality and safety of patient care, the aim is to automate the classification of screening mammograms using Machine Learning techniques. The development will be carried out in the MATLAB environment [25]. In a near future, the developed system can be transferred to other environments more suitable for daily use, such as a windows application developed in C++ and compiled for distribution and use by personnel not trained in programming. This system can reduce costs and avoid unnecessary medical procedures. Ultimately, the proposed system would act as an initial step for the diagnosis of breast cancer.

The dataset used to feed the neural network consists of a total of 54707 images, provided by the Radiological Society of North America (RSNA) through the Kaggle platform [26].

This group of data includes mammograms made according to 4 different types of projections: cranio-caudal (CC), medio-lateral oblique (MLO), medio-lateral (ML) and latero-medial (LM). There are only mammograms with cancer in the first two groups (CC and MLO) so the ML and LM types (very rare in the database) are ignored. The CC projection is obtained by applying compression from the upper part of the breast that is resting on the surface of the detector, with the beam of rays being perpendicular to the ground. In the case of the MLO, the rays are almost parallel to the ground with a slight inclination. In this case, sometimes, an additional X-ray image is obtained to cover the axillary region. The ML and LM projections are similar to the MLO, with the difference that the rays go completely parallel to the floor, outwards from the center of the chest (ML) or inwards from the outside to the center (LM).

### 2.1. Preprocessing

Before proceeding to the design of the CNN, the preparation of the images is necessary to adjust all of them to a common format and characteristics. Starting from a dataset of images classified into folders by patient number and a csv text file with data about the images. Each folder contains at least 4 images, two projections for the left breast and two for the right one (CC & MLO for each). However, sometimes other types of projections are included or some of the previous ones are repeated, so there are certain folders that contain more images. One of the reasons could be the correct display of certain areas. The data provided by the csv file are, among others, the patient and image codes, what type of projection it is and whether it is positive or not for malignant cancer. With this data, mammograms are grouped into 8 types: CChealthy, MLOhealthy, LMhealthy, MLhealthy, CCsick, MLOsick, and LMsick; although the last two groups will be empty. Next, to make the dataset as homogeneous as possible a preprocessing stage is applied to all images. Mammograms with image information on the right are flipped so that interest part is always on the left, contrast is modified depending on whether the breast is dense or fatty, size is made uniform (256×256), pixels are normalized to real values between 0 (black) and 1 (white). An example of this process can be seen in Figure 2. Note that the detection of the side of interest (left or right) and the classification as fatty/dense are automatically done using classical image processing techniques.

**Figure 2.**
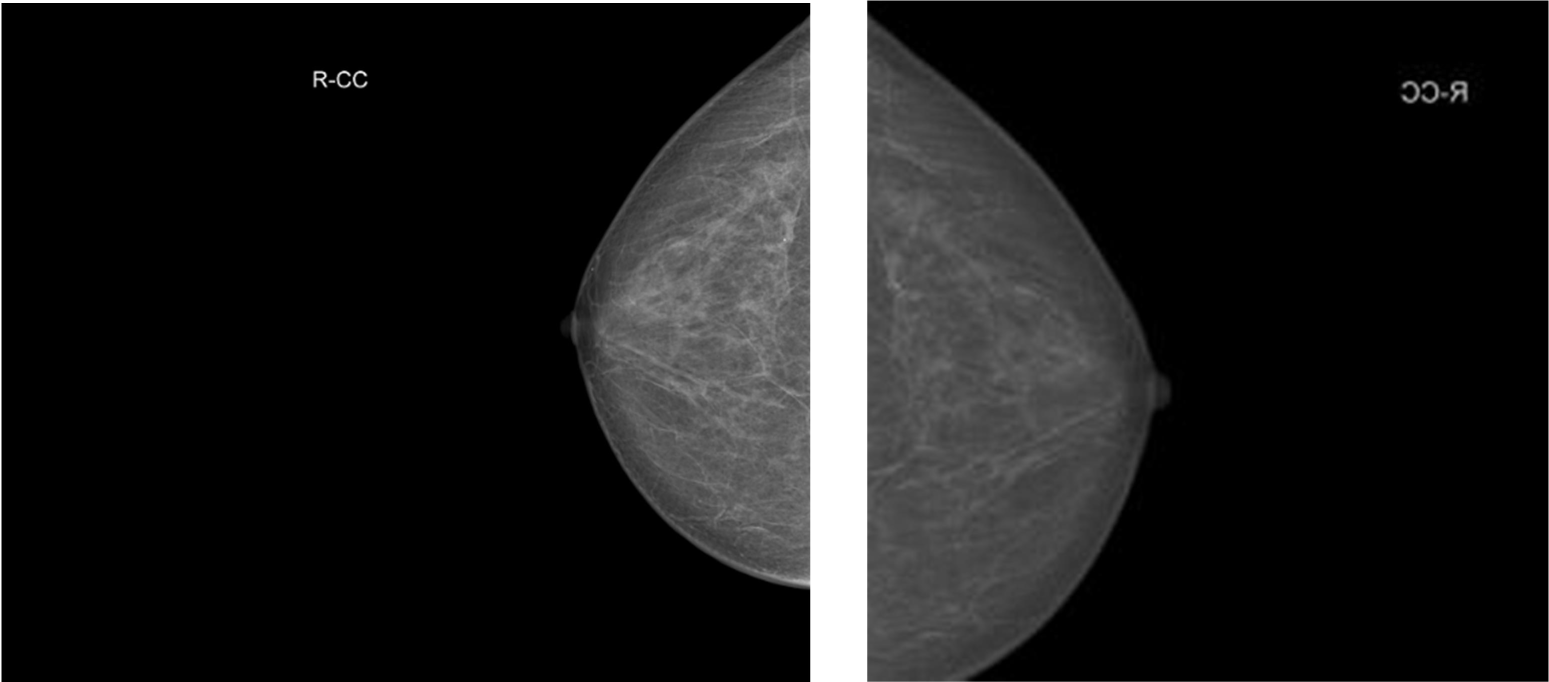
CC (right breast) mammogram before (left) and after (right) processing.

In addition, another type of processing (Preprocessing 2) is tested. The main difference is that one more step is added: the program detects the largest object in the mammogram (i.e. the breast) and image is cropped to the bounding box of this object, then cropped image is resized to the same final size: 256×256. This allows that the region of interests occupies as much space as possible, without varying the texture information. The price is some deformation that might not be a problem for Machine Learning algorithms since the discriminant information is in the texture.

**Figure 2.**
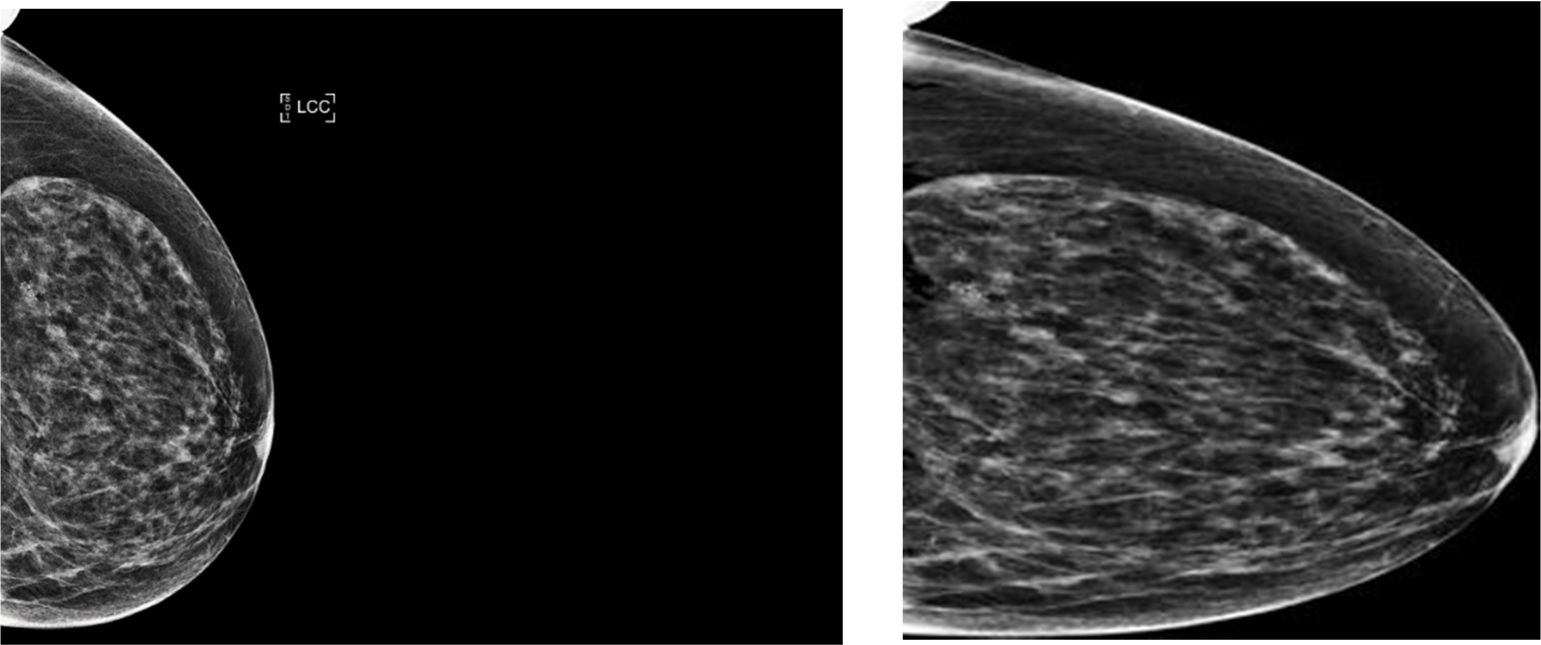
Mammogram (CC of left breast) before (left) and after (right) processing with method 2.

### 2.2. Solution to class unbalance

Initially, of the 54707 starting images, there are 566 mammograms with cancer visible in the CC projection and 590 with cancer visible in the MLO projection. So there is clearly an important unbalance between the healthy and sick classes.

As a solution to this mismatch, several methods are proposed. First, the majority class is under-sampled, keeping a total of 1132 CC projection images (566 from each class) and 1180 images for the MLO projection (590 from each class).

Other method is augmenting the minority classed using a function called **`randReplicateFiles`** (downloaded from Matlab Central [27]), which balances both groups of images, replicating those of the minority group. In this case, we added new images to the “Healthy” folder until we tripled the initial number, obtaining 3396 CC images (1698 per class) and 3540 MLO images (1770 per class).

Another oversampling option tested is to create new images that are very similar visually but really different using a new function (**`ForgeImage`**). **`ForgeImage`** is a function that was developed for other purposes (in the field of image forensics and other computer security applications [28]) and modifies an image slightly by adding random noise and performing slight geometric modifications. The intensity of these transformations can be adjusted so that the visual effect is practically imperceptible.

### 2.3. CNN Design

The performance of CNN’s depends, to a large extent, on their architecture; i.e. the number and type of layers and the number of filters. By network design we mean the decision on the most appropriate number of filtering layers, the size and number of filters in each one, the processing points where normalizations are to be made, the type of activation functions to be applied, the type and size of the decimation layers and the number of “fully connected” (MLP) layers. In the work of designing the CNN, various configurations are tried to obtain the best results. In general, the network is made up of convolutional and subsampling (or decimated) layers, which are grouped into modules or blocks. Through the application of convolutional filters, high/medium level information is extracted, which the network will interpret as characteristics. As the number of filters increases, this information becomes more refined.

The basic layers of a CNN are the input layer, the convolutional layer, the clustering layer and the fully-connected layer.

First, the input layer feeds 2D images to the neural network and applies data normalization. The convolutional layer is the central building block of a CNN, where most of the calculations happen. Groups of pixels close to each other are taken and mathematically operated (scalar product) against a small matrix (kernel). This kernel loops through the entire image, generating a new output image, which will feed the next layer of neurons. Actually, not just one kernel is applied, but a group of them; so as many new images are computed as the number of filters in the layer. These new images are “showing” certain characteristics of the original image, which will help to distinguish one group of images from another.

Two processes are then typically used: normalization and applying an activation function (the most common is the so-called ReLU function). A pooling layer immediately appears that reduces the amount of data before doing a new convolution in order to remove redundant information. One of the most used methods is Max-Pooling, which consists of, for example, if we have a 2×2 matrix size, for each group of 4 pixels in the previously generated image, the highest value among those 4 is extracted and so on with each group of pixels, until we get a decimated image half the width and half the height.

Finally, in the fully connected layer (**fullyConnectedLayer**) each of the outputs of the previous layer is connected to this one with a certain weight. This stage is a classical Multi-Layer Perceptron (MLP).

After the convolutional network is defined, it can be trained, adjusting each of the numerical parameters (filter coefficients and MLP weights) with the aim of minimizing the error between the obtained and the desired output. For this task, a **sgdm** (Stochastic Gradient Descent with Momentum) training engine is used. The stochastic gradient descent algorithm updates the parameters (weights and biases) to minimize the error function by taking small steps in the direction of the negative gradient of the loss function. Several trials are performed at this point for different combinations. First, images are pre-processed using two different methods (methods 1 and 2 described above). Second, two types of oversampling methods are tested (**`randReplicateFiles`** and **`ForgeImage`**). For each option CC and MLO projection images are tested independently and also mixing images of both types together (**MIX**).

After training the networks, tests are carried out to check if the operation is as desired using a different set of data (to avoid the problem of overfitting). In general, the CNN trains with a percentage of between 80% and 90% of the total mammograms.

### 2.4. Assesment measurements

After ANN’s have been trained, some parameters are calculated to quantify the quality of each of the networks in order to compare them. These parameters are: *Accuracy*, *Confusion*-*Matrix*, *Precision*, sensititvity (*Recall*) and the parameter F (*F_1_-score*) for each of the trainings. In this way, conclusions can be drawn and improvements can be researched.

The first parameter that the networks return is *Accuracy*, the percentage of samples that the model has classified correctly. This indicator is not highly recommended for unbalanced data sets because the network will tend to classify all cases as belonging to the majority class.

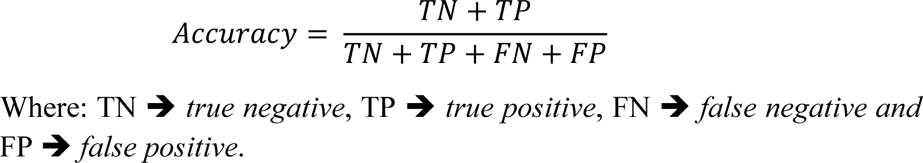

Therefore, one measurement that clearly shows whether the network is confusing classes when classifying images is the *Confusion-Matrix*. Each column of the matrix represents the number of predictions for each class, while each row represents the actual (target) classes.

|  |  |
| --- | --- |
| TN | FP |
| FN | TP |

From this matrix, other indicators can be obtained such as: *Precision* (probability that a true classification is really true) and sensitivity (or *Recall*: the probability that a true case is not missed) are also obtained. Finally, the F1-score measurement is the harmonic average between the two previous factors.

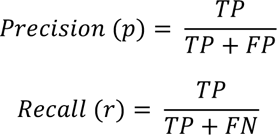

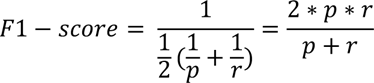

**Figure 3.**
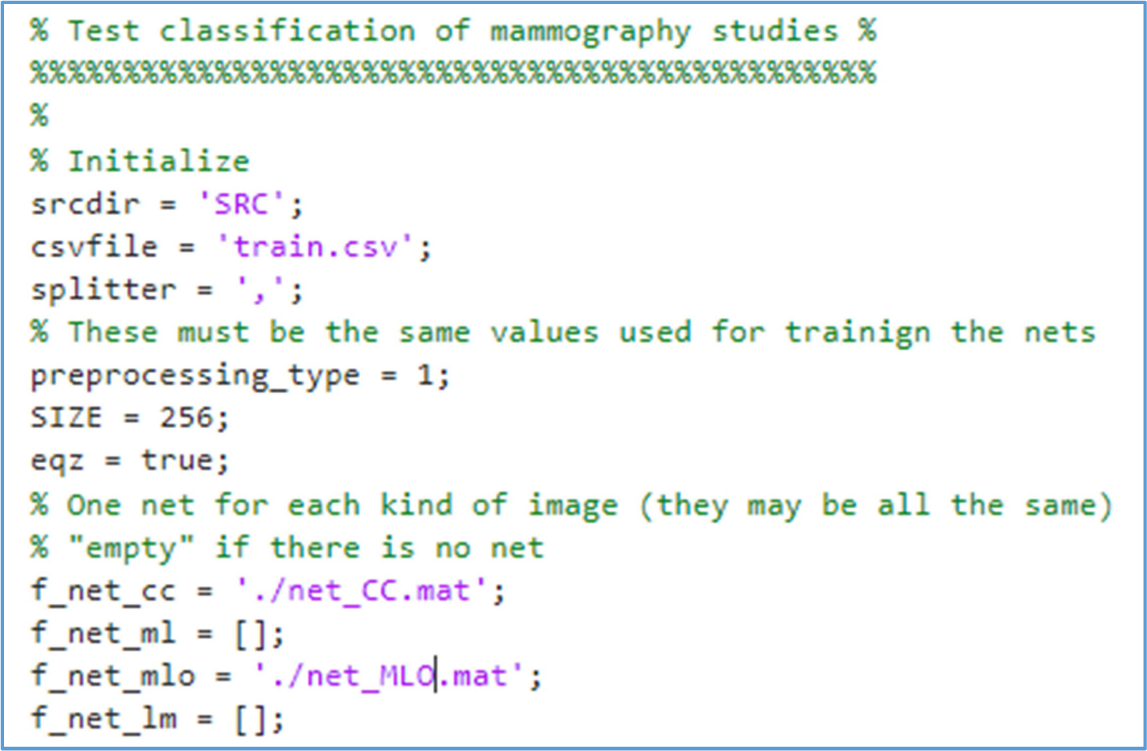
Global test inputs.

### 2.5. Global test

After training the network and validating it with a reduced group of images, to extract the results described above, the global test is carried out. This consists of a MATLAB program in which a larger group of images is selected from the database with which it was trained, to classify images and patients according to whether or not they have a cancer positive diagnosis. As inputs to this program are the folders that belong to each patient (SRC general folder), with their corresponding images (radiographies), the previously trained networks (**`net_CC.mat`** and **`net_MLO.mat`**, in this case) and the **`.csv`** file (**`train.csv`**), as it can be seen in Figure 4. The outputs are the values of *Precision*, *Recall*, *F_1_-score*, *Accuracy* and *Confusion-Matrix*. Two sets of these parameters are computed: image-wise and patient-wise. First, the program treats each patient as two independent entities (two breasts), considering as independent the images from the right and the left breast; since the cancer can be in the right one, in the left one or in both. Second, when an image is classified as a sick breast, that patient will be classified as sick. The program directly reads the images in DICOM format and then applies preprocessing 1 or 2, depending on which was used at network training; finally, the network categorizes it as “sick” or “healthy”.

**Figure 4.**
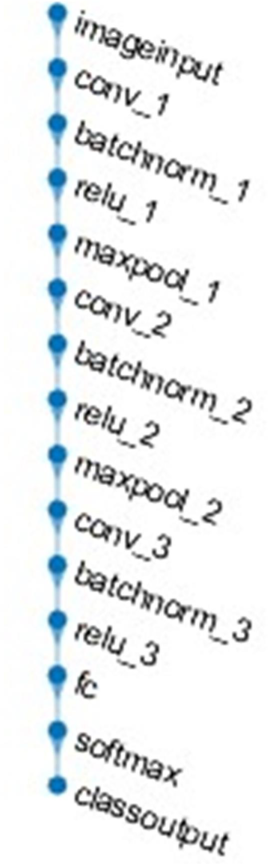
Architecture of the convolutional neural network used.

This global test is important to see the results obtained with images that have not been used before. In addition, a more realistic view of the process is obtained, testing with many real world images that present a strong unbalancing between classes, just as in the real world.

## 3. Results and discussion

The algorithms proposed in this study are implemented in MATLAB (R2022b). As previously mentioned, the methodology involves multiple steps that include image processing techniques, CNN training and the implementation of techniques that balance both classes. The images are pre-processed according to two different methods, and then the CNN is trained with the resulting dataset (once balanced)

To define neural network configuration in MATLAB, the following “layer-defining” instructions are used:

- **imageInputLayer**: two input layer dimensions were tested: 256×256 and 512×512.
- **`convolution2dLayer`**: 3×3 and 5×5 filter sizes are used, in 3 convolutional layers. Different settings on the number of filters are tried, including 8, 16, 32 and 8, 8, 16. The padding parameter is set as “same”, which ensures for each that output is of the same size as the input.
- **`batchNormalizationLayer`**: batch normalization layer, speeds up network training and reduces initial sensitivity. It is located between the convolutional and activation layers.
- **`reluLayer`**: in this layer, every input element less than zero is set to zero. It is an activation function mathematically equal to:

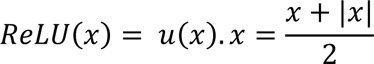
- **maxPooling2dLayer**: a size of 4×4 is set after the first convolutional layer and 3×3 after the second, with a step size of 4×4 and 3×3 respectively.
- **`fullyConnectedLayer`**: layer with an output size of 2. All neurons in a fully connected layer are connected to neurons in the previous layer, combining all features learned by previous layers to identify patterns.
- **`softMaxLayer`**: nonlinear activation function (*softmax*) to compute the output values. Softmax produces “probability like” scores that add up to 1.0.
- **classificationLayer**: output layer that computes the classification result (selects maximum activation).

The network architecture displayed by MATLAB tools can be seen in Figure 5:

**Figure 5.**
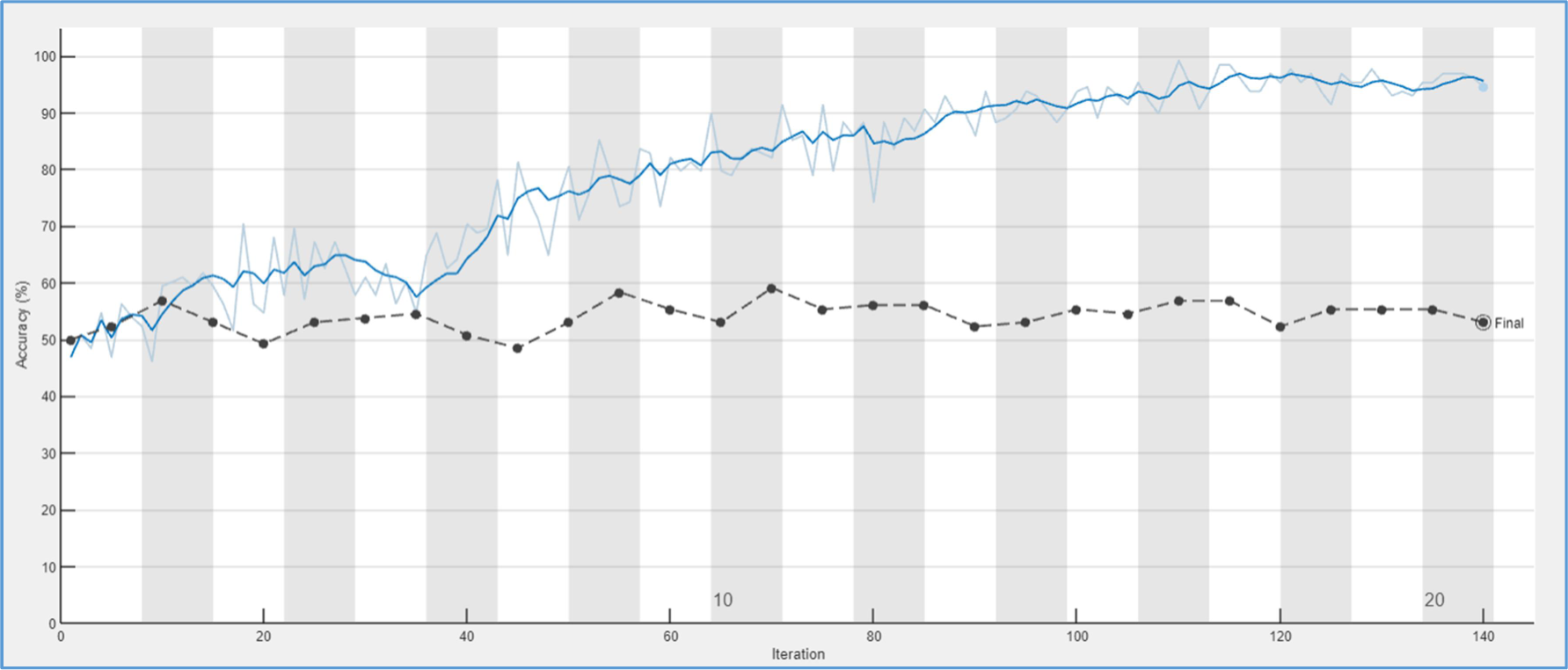
Accuracy curves obtained training the network without oversampling, mammograms with CC projection.

**Figure 7.**
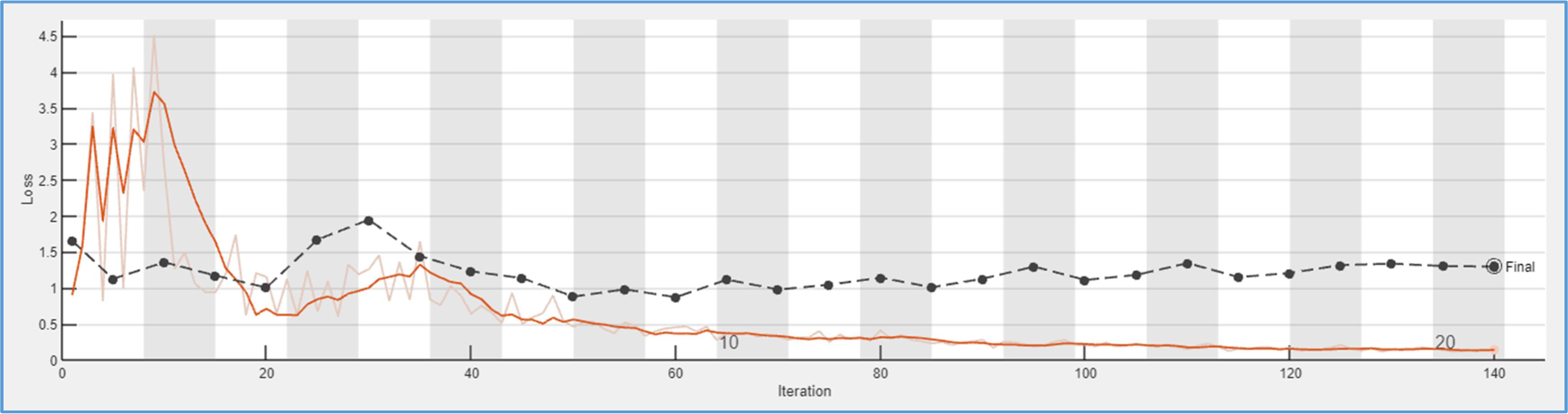
Mean square error obtained training the network without oversampling, mammograms with CC projection.

### 3.1. Network training without oversampling and Preprocessing number 1

The first test that was carried out consisted of training the network with the CC projection mammograms. 1132 images were chosen, i.e. 566 healthy and 566 with cancer. 566 is the total number of breast radiographs of type CC with disease. 500 images from each class were chosen for training and the rest for further validation. The maximum number of executions of the algorithm is fixed at 20 epochs. An epoch consists of training once with the entire data set, and each epoch is divided into iterations. An iteration (or **`miniBatch`**) consists of running for M samples and updating the weights, M size is 128 by default. A validation frequency of 5 means that, every 5 iterations, a test is made with the validation samples, obtaining *Accuracy* and mean squared error (MSE) values, building a curve that allows to check the progress of the training process. In addition, the number of filters for each convolutional layer was 8 in the first one, 16 in the second one and 32 in the third one. The results obtained can be seen in Figures 6 and 7.

**Figure 6.**
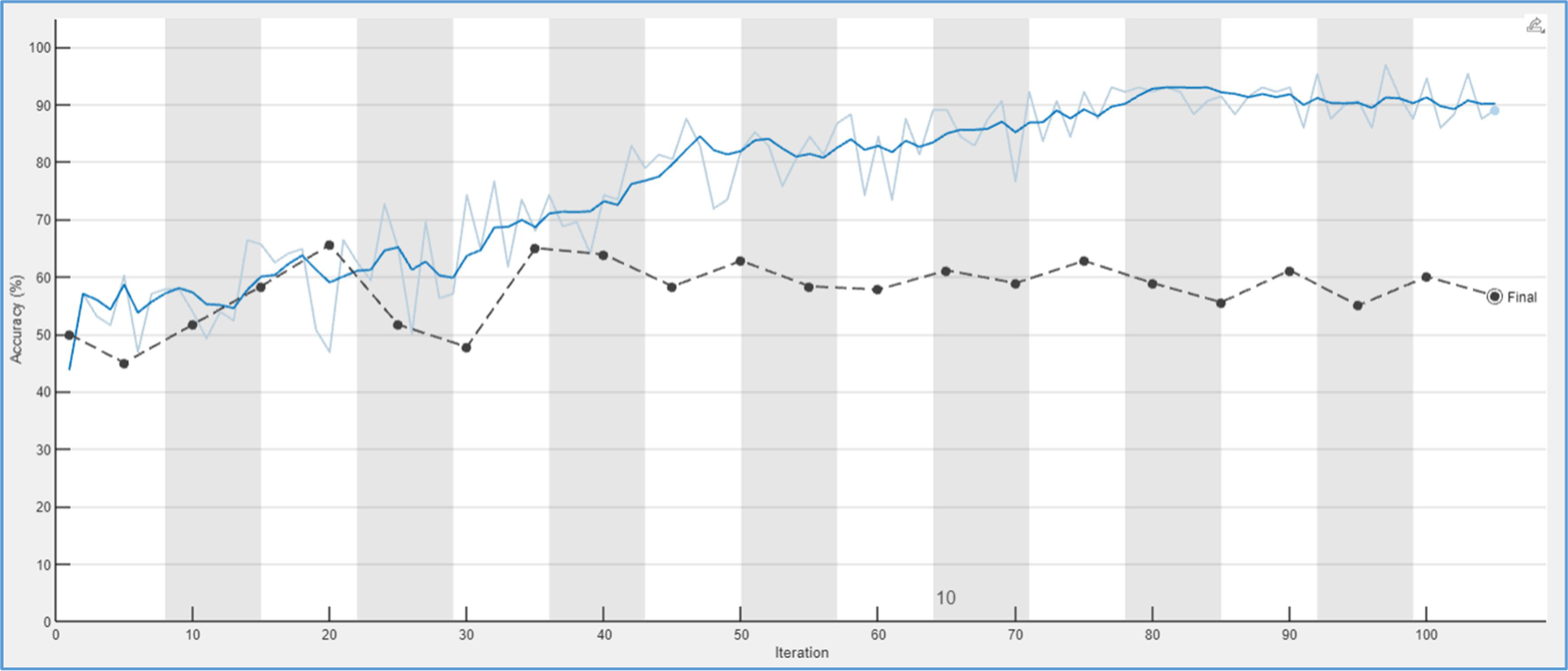
Accuracy curves obtained training the network without oversampling, mammograms with MLO projection.

**Figure 7.**
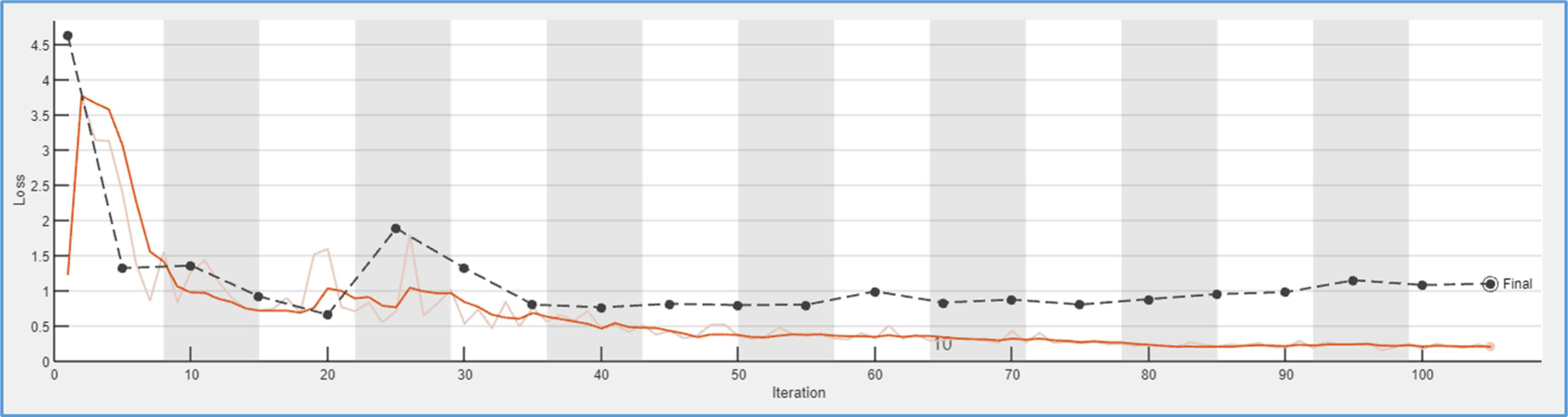
MSE obtained training the network without oversampling, mammograms with MLO projection.

At first glance, it can be seen that although the network is able to train correctly and obtain accuracy values close to 95%, the validation curve is around 53%. This fact indicates that more training images are needed (high *Accuracy* is due to overfitting). For mean square error (MSE) values, the conclusion is basically the same.

Testing with the group of mammograms with MLO projection, 590 healthy and 590 sick images were selected. The results obtained can be seen in Figures 8 and 9. In this case, 15 epochs were chosen and the accuracy of the final validation is around 56%.

**Figure 8.**
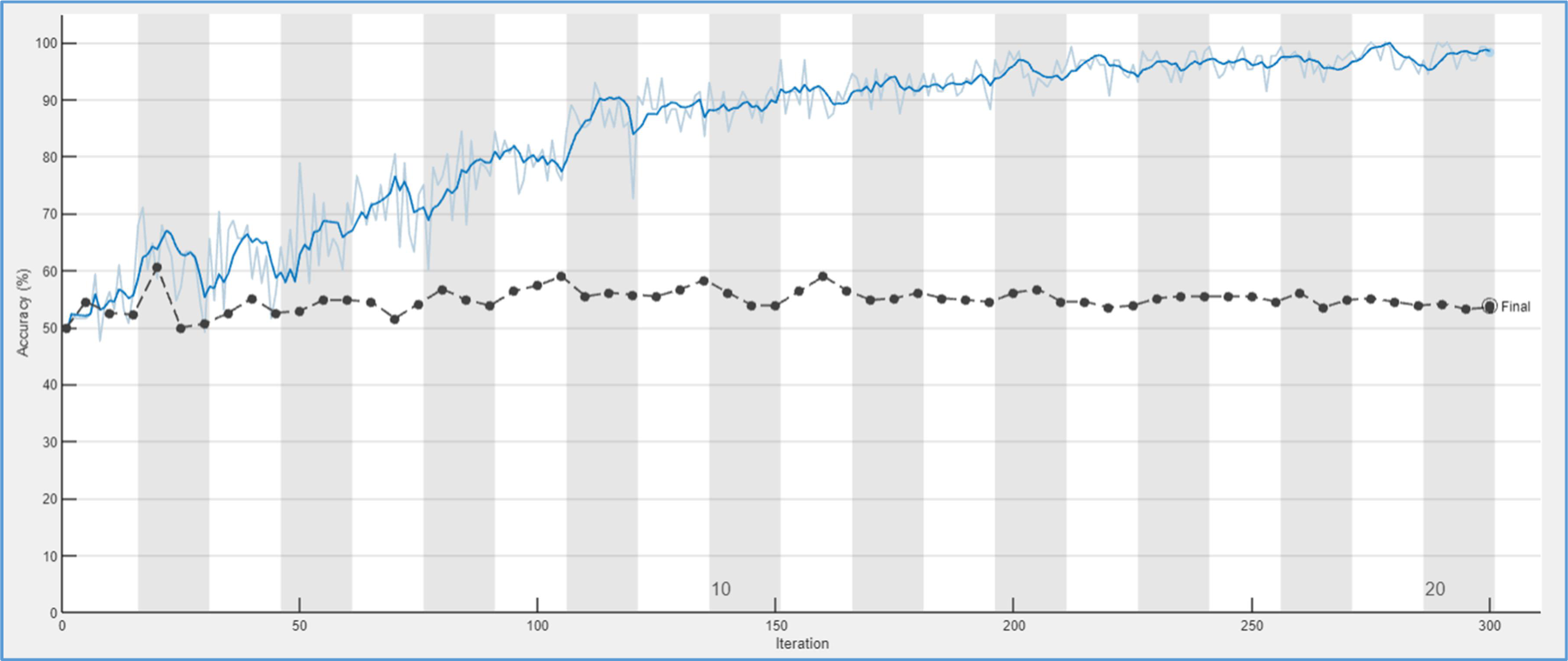
Accuracy curves obtained training the network without oversampling, mixing CC & MLO mammograms.

**Figure 9.**
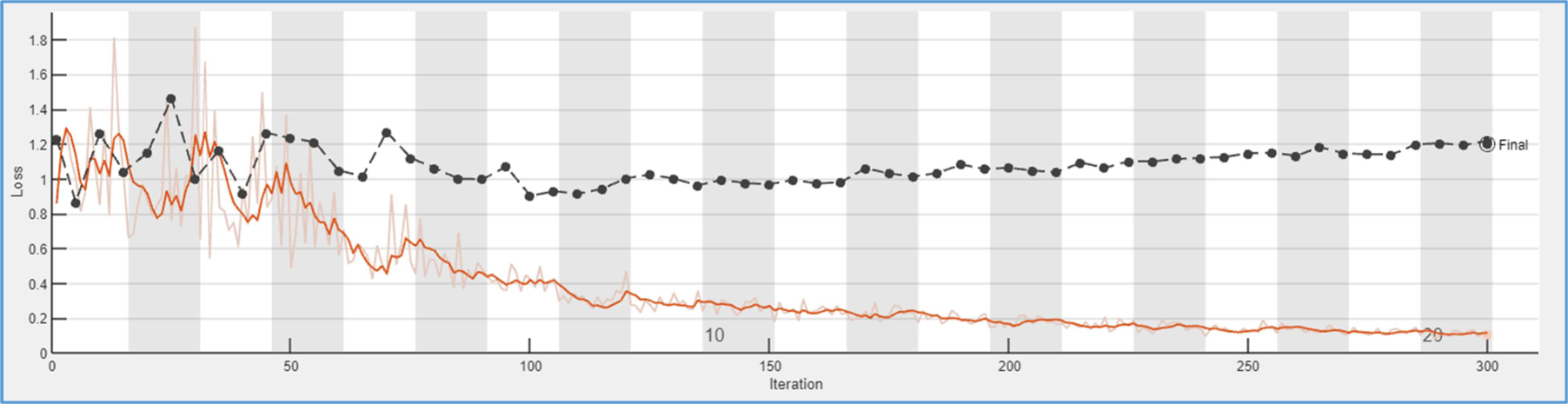
MSE curves obtained training the network without oversampling, mixing CC & MLO mammograms.

Very similar results to the previous ones are obtained, which indicates that strong class unbalance makes that training set (without oversampling) is enormously reduced causing a bad generalization. Next try was to study the influence on *Accuracy* by mixing both types of projections. So the network was trained with 566 images with CC projection and 590 with MLO in both classes (healthy and sick). The results obtained can be seen in Figures 10 and 11.

**Figure 10.**
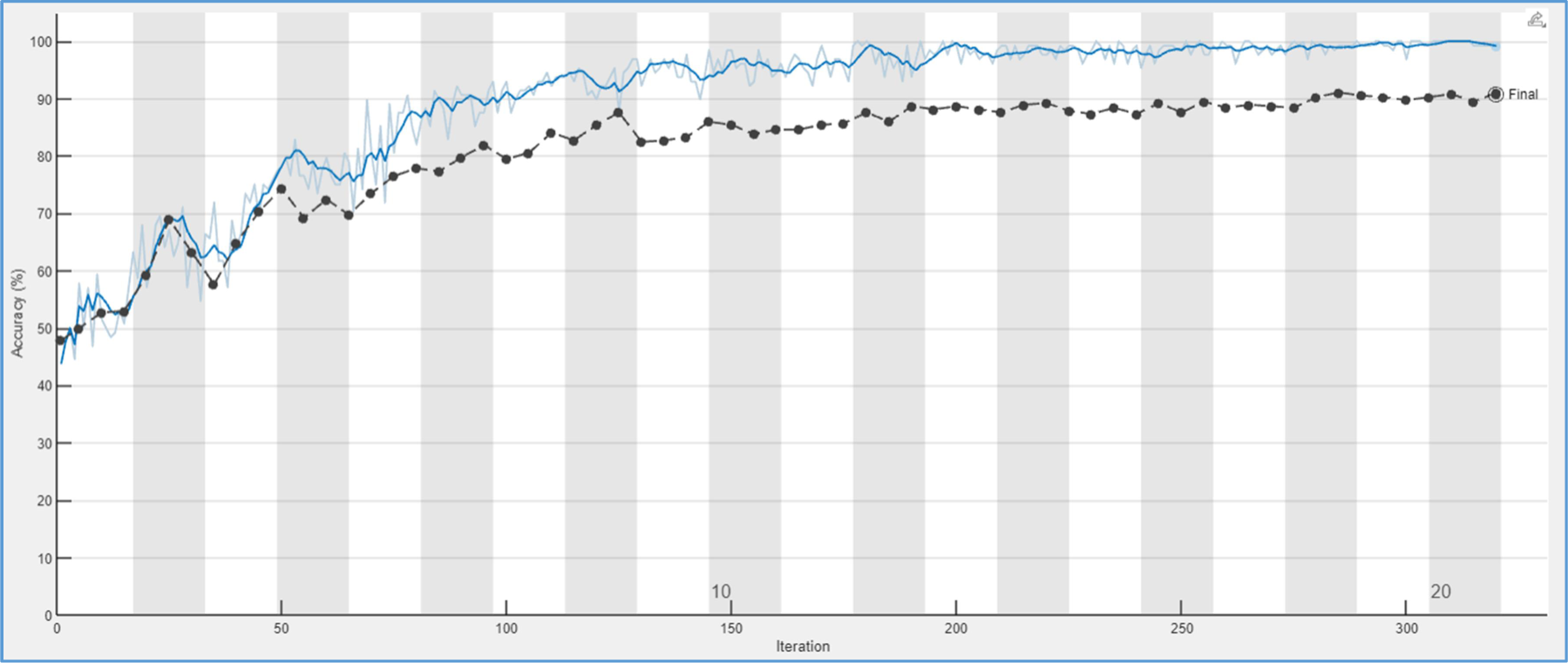
Accuracy curves obtained training the network oversampling the sick images (replication), mammograms with CC projection.

**Figure 11.**
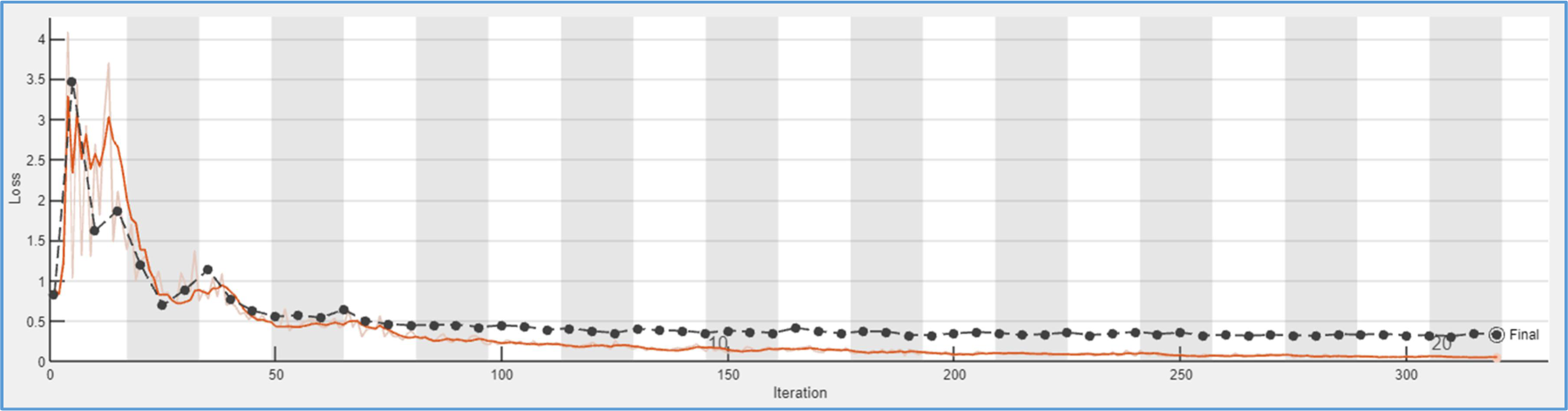
MSE curves obtained training the network oversampling the sick images (replication), mammograms with CC projection.

*Accuracy* remains similar to the previous tests, and the loss function increases slightly, around 0.2. These results make us think that there are not too many differences in training the network by combining both projections or separately.

### 3.2. Network training with oversampling (replication: randReplicateFiles)

To overcome the problems described in the previous section, a solution can be the use of oversampling. The first oversampling method used is simply file replication, using the function **`randReplicateFiles`** (from MATLAB central).

A dataset was created consisting of a folder with twice as many healthy as diseased images extracted from the previously processed database. Of the 26199 radiographs without breast cancer, 566 more are chosen. The dataset was then balanced applying the replication function to the sick images. The network was trained using 85% of the training images, resulting in the results shown below in Figures 12 and 13. With oversampling, accuracy goes from approximately 53% to 90% and the loss function (MSE) is reduced by almost 1 unit.

**Figure 12.**
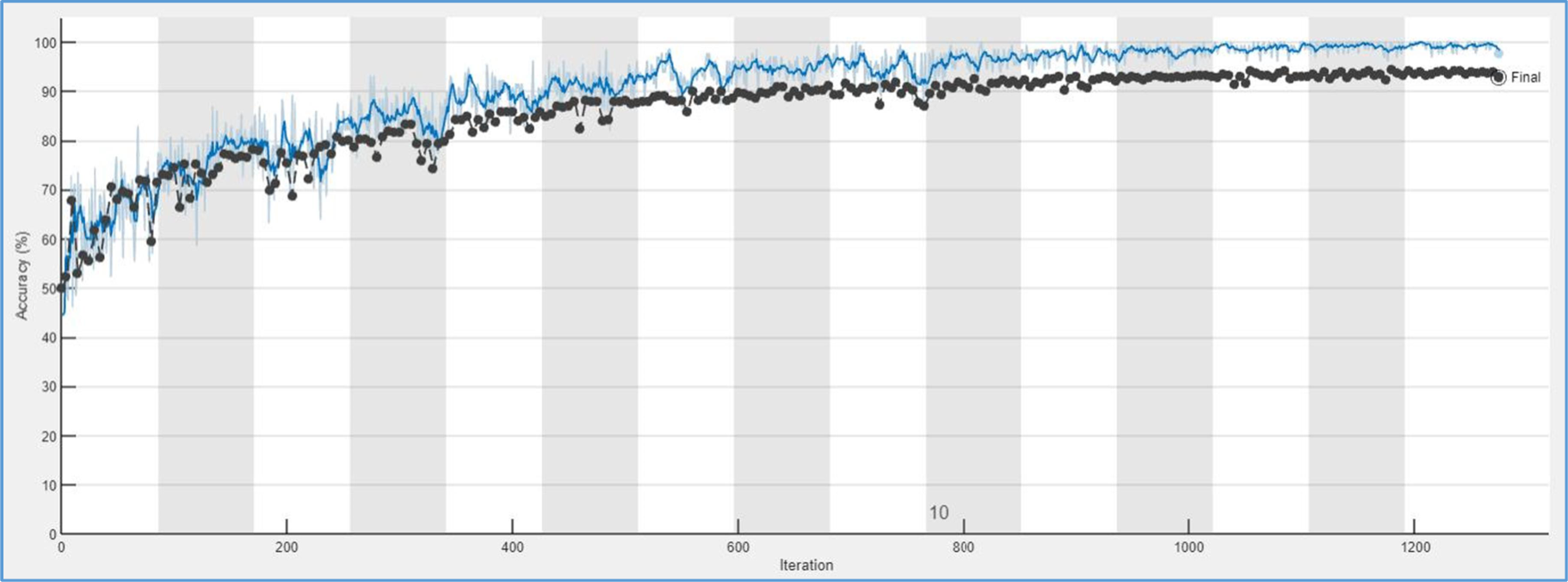
Training with extended dataset: *Accuracy* curves.

**Figure 13.**
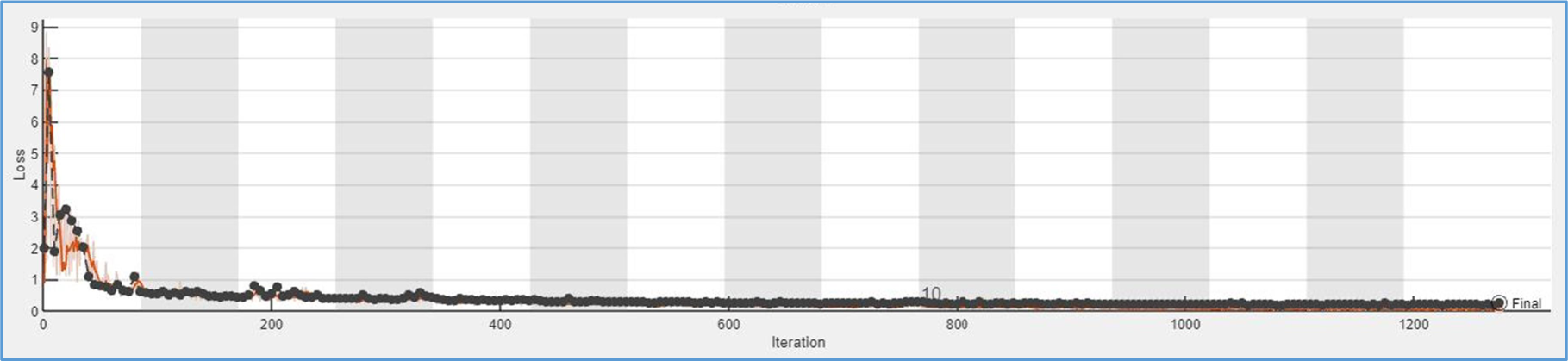
Training with extended dataset: MSE (mean square error).

The obtained results confirm the idea that to achieve optimal training, it will be necessary to increase the minority class. In light of this, it was decided to also try tripling and quadrupling the images of the minority class. To compare the experiments, the *Accuracy*, *Precision*, *Recall*, and *F1-score* were calculated for each case using the formulas presented in the previous section. As we increased the size of the dataset, accuracy improved. However, the results obtained from tripling the images of the minority class were better than those from quadrupling them, possibly because excessive repetition of samples can cause a form of overfitting. See results in table 1.

**Table 1.** *Accuracy, Precision, Recall, and F1-score* results for different oversampling rates in the minority class for(CC images).

|  | *Accuracy (%)* | *Precision (%)* | *Recall (%)* | *F1-score (%)* |
| --- | --- | --- | --- | --- |
| No oversampling | 57,53 | 58,06 | 54,22 | 56,07 |
| Duplicating | 88,05 | 84,96 | 92,48 | 88,53 |
| Tripling | 93,15 | 89,49 | 97,78 | 93,45 |
| Cuadruplingr | 91,32 | 85,29 | 96,99 | 90,76 |

Oversampling was also applied to the case of mammograms with MLO projection and to the joint dataset of both types mixed (MIX). In this case, the number of images was tripled in both cases, and the obtained results can be seen in Table 2 below.

**Table 2.** *Accuracy, Precision, Recall, and F1-score* results for CC, MLO, and MIX projections, tripling the minority class.

|  | *Accuracy (%)* | *Precision (%)* | *Recall (%)* | *F1-score (%)* |
| --- | --- | --- | --- | --- |
| CC | 89,63 | 84,74 | 96,67 | 90,31 |
| MLO | 91,51 | 89,29 | 94,34 | 91,74 |
| MIX | 89,25 | 85,35 | 94.77 | 89,81 |

Next, a small test was conducted to check the importance of the number of filters in the convolutional layer, and the following results were obtained (Table 3). Oversampling was used to duplicate the images of the minority class in the 3 experiments, and 85% of the images were used for training.

**Table 3.** Comparison of *Accuracy, Precision, Recall, and F1-score* with different number of filters (CC projection).

|  | *Accuracy (%)* | *Precision (%)* | *Recall (%)* | *F1-score (%)* |
| --- | --- | --- | --- | --- |
| 8, 8, 16 | 86,76 | 83,01 | 82,43 | 87,47 |
| 7, 14, 28 | 89,71 | 83,55 | 85,29 | 89,23 |
| 8, 16, 32 | 88,05 | 84,96 | 92,48 | 88,53 |

In the first case, the figures of merit are generally lower because the number of applied filters is also lower, resulting in fewer features extracted. In the following two experiments, the *Accuracy* is similar. Since *Recall* is one of the most important variables in the medical field, it is chosen for selecting the best method. The *Recall* in the third method increases by approximately 7% compared to the second. Therefore, the number of filters is 8, 16, and 32 in each successive convolutional layer.

As mentioned earlier, the images have a size of 256×256 pixels. It was decided to investigate if resolution could affect network performance. Therefore, a small experiment was conducted where CC projection radiographs of size 512×512 pixels were selected and duplicated using the previous method. Additionally, the mini-batch size was changed from 128 (default) to 64 to avoid memory overflow errors. The results obtained were as follows: *Accuracy*: 87.06%, *Precision*: 86.21%, *Recall*: 88.24%, and *F1-score*: 87.21%. Compared to the lower resolution images, there were no significant improvements in training, so it was decided to continue with the lower resolution images, which also allow for faster processing times.

### 3.3. Network training with oversampling (modification: **ForgeImage**)

After performing all the tests applying the first type of oversampling, and to research whether this network can be improved; the following method: **`ForgeImage`**, is applied and the obtained results are compared. In this case, the images are processed and then put into the network training set. For the ‘sick’ images folder, for each ‘real’ image, one (or two) ‘fake’ image(s) is(are) created. In this way, the number of mammograms in the minority group is doubled or tripled. The obtained results, compared to the previously applied oversampling, are shown in Tables 4 and 5.

**Table 4.** *Accuracy, Precision, Recall, and F1-score* results for different oversampling options (CC images).

|  | *Oversampling type* | *Accuracy (%)* | *Precision (%)* | *Recall (%)* | *F1-score (%)* |
| --- | --- | --- | --- | --- | --- |
| CC | *randReplicateFiles (X2)* | 88,05 | 84,96 | 92,48 | 88,53 |
| CC | *ForgeImage (X2)* | 71,97 | 71,97 | 71,97 | 71,97 |
| CC | *randReplicateFiles (X3)* | 93,15 | 89,49 | 97,78 | 93,45 |
| CC | *ForgeImage (X3)* | 82,06 | 78,84 | 75,25 | 77 |

**Table 5.** *Accuracy, Precision, Recall, and F1-score* results for different oversampling options (MLO images).

|  | *Oversampling type* | *Accuracy (%)* | *Precisión (%)* | *Recall (%)* | *F1-score (%)* |
| --- | --- | --- | --- | --- | --- |
| MLO | *randReplicateFiles (X2)* | 89,83 | 90,29 | 89,27 | 89,77 |
| MLO | *ForgeImage (X2)* | 76,54 | 80,53 | 70,00 | 74,90 |
| MLO | *randReplicateFiles (X3)* | 91,51 | 89,29 | 94,34 | 91,74 |
| MLO | *ForgeImage (X3)* | 81,14 | 85,49 | 75,00 | 79,90 |

As can be seen at first glance, the results obtained by applying the second type of oversampling are worse than the previous ones, decreasing the values by 10 or 15%. This may be because this type of oversampling creates small (almost non visible) errors but that can be of importance for cancer detection.

The data augmentation method provided by Matlab [29] was also applied. This tool (**`imageDataAugmenter`**) allows the creation of new images by rotating, cropping, and translating them so that the network trains better. However, in this specific case, it did not work because although the validation curve ‘sticks’ closer to the training curve, the latter stalls at 72%. This method may work well for augmenting images when dealing with ordinary photographs of common objects. In this other problem the network needs to recognize objects even if they are in different positions, have different sizes, and colors… In this problem the object is always in, approximately, the same position and it is also of approximately the same size.

### 3.4. Network training with Preprocessing number 2

After training the network with the images converted using Processing 1 and applying both types of oversampling to these images, the second type of processing is executed to see if the results can be improved. In this case, as mentioned earlier in the methods and materials section, the images are deformed in such a way that the uninteresting black background information is cropped out, and the texture of the breast, which provides information about cancer, is expanded.

Selecting the best parameters from the previous experiments, namely, number of filters in each convolutional layer: 8, 16, and 32; 20 epochs; and a validation frequency of 5. Additionally, the number of training images chosen is 85% of the total.

The obtained results are summarized in Table 6.

**Table 6.** *Accuracy, Precision, Recall, and F1-score* results for mammographies pre-processed using the second methods and for different *oversampling options*.

| *Projection type* | *Oversampling type* | *Accuracy (%)* | *Precision (%)* | *Recall (%)* | *F1-score (%)* |
| --- | --- | --- | --- | --- | --- |
| CC | None | 60,61 | 62,96 | 51,52 | 56,67 |
| MLO | None | 56,11 | 55,67 | 60,00 | 57,75 |
| MIX | None | 53,85 | 53,49 | 58,97 | 56,10 |
| CC | *randReplicateFiles* (X3) | 91,96 | 89,93 | 94,51 | 92,16 |
| MLO | *randReplicateFiles* (X3) | 91,51 | 90,74 | 92,45 | 91,59 |
| MIX | *randReplicateFiles* (X2) | 83,43 | 79,00 | 91,07 | 84,61 |
| CC | *ForgeImage* (X2) | 75,38 | 72,79 | 81,06 | 76,70 |
| CC | *ForgeImage* (X3) | 80,30 | 80,30 | 80,30 | 80,30 |
| MLO | *ForgeImage* (X2) | 77,50 | 76,19 | 80,00 | 78,05 |
| MLO | *ForgeImage* (X3) | 79,00 | 77,10 | 82,50 | 79,71 |

As it can be seen, the overall results are worse compared to Tables 4 and 5, meaning that the most effective approach is still training with images pre-processed using method 1 and the randReplicateFiles type of oversampling. For example, the best results achieved with preprocessing 1 are as follows: *Accuracy*: 93.15; *Precision*: 89.49; *Recall*: 97.78; and *F1-score*: 93.45. When comparing these results with those of pre-processing 2 for the same training, the *Accuracy* and *F1-score* decrease slightly, whereas *Precision* increases almost insignificantly. Key fact is that *Recall* (the most important parameter in a diagnostic system), decreases by 3%. This may be due to the fact that the size of the breasts among patients varies greatly, whereas the resulting breast with pre-processing 2 will have always a similar size, causing certain characteristics of the tumor masses to be affected and not being optimally recognized by the CNN.

Some main characteristics of benign breasts are that they generally have low density, well-defined margins, and a cover made of fat over the lesions. While malignant ones usually have an irregular shape, no symmetry, and no fatty covering. On the other hand, malignant micro-calcifications, as mentioned in the introduction, tend to be small, of different sizes, and have irregular edges, whereas benign ones are large, homogeneous, and round [30]. For all these reasons, deforming the images may cause the differences between malignant and benign to become less prominent, and consequently, the network may not be able to work correctly.

Finally, seeing the generally satisfactory results of the model, a new MATLAB program is coded to classify a larger group of images that has not been balanced. This is interesting to confront the system with the real world. As mentioned earlier, the results are offered with respect to the images and to the patients (each breast can be seen as an independent patient or a patient can be classified as sick if at least one breast is classified so). Two tests have been performed: first, a different network is used for each type of projection and, second, a CC trained network is used to classify all images. There are clear differences between the obtained results, so it can be assumed that classification worsens when only one type of pre-trained network is used. The *Recall* in the case of results obtained by image goes from 76% to 90% and by patient from 94% to 97%. Therefore, it can be concluded that there are slight differences between the features the network extracts from the CC and MLO projections.

Next, a new test is carried out where the network introduced was trained without oversampling. In this case, *Recall* drops to 69% for images and 88% for patients. As previously demonstrated, oversampling of the minority class is necessary for the classification program to achieve the best possible results.

Subsequently, the network trained with mammograms with both CC and MLO projections, treated with preprocessing 1, is tested. Significant results are obtained as the *Recall*, i.e., the program’s sensitivity, increases considerably. Values of 97% are obtained in the case of image classification, and 100% in the case of patients, thus the number of false negatives obtained for this case was zero.

To further improve these results and obtain better *Precision* and *F_1_-score* values, the database is expanded with mammograms extracted from a new Kaggle dataset [31]. These new mammograms are processed differently as they are separated into 4 independent folders according to whether they contain cancer or not and whether they are breasts with implants or not (’cancer_negative’, ‘cancer_positive’, ‘implant_cancer_negative’, and ‘implant_cancer_positive’). Only the folders positive for cancer are selected, where the images are mixed (mammograms with CC and MLO projection), and the resolution is changed from 512×512 to the previous one used, i.e., 256×256. The aim is to train the network with both the old and new sick mammograms, and also to increase the number of healthy images. A total of 984 mixed projection images are obtained, which, added to the previous 1156, makes a total of 2140.

Applying a randReplicateFiles type of oversampling to triple the number of sick breast radiographs (6420), the network is trained, and the graphs shown in Figures 14 and 15 are obtained.

These results are very promising as both the training and validation curves approach each other and are very close to optimal limits. By introducing this new network into the overall program, the results shown in Table 7 are obtained.

**Table 7.** Results for 40% of whole dataset.

|  | Per image | Per patient |
| --- | --- | --- |
| *TN* | 16497 | 5487 |
| *TP* | 429 | 198 |
| *FN* | 32 | 0 |
| *FP* | 4840 | 3831 |
| *Precision* | 8% | 5% |
| *Recall* | 93% | 100% |
| *F<sub>1</sub>-score* | 15% | 10% |
| *Accuracy* | 78% | 60% |

In view of these results, it can be assumed that this program is very thorough, meaning it is very unlikely to incorrectly classify a real cancer as negative. This is very interesting in this specific medical application because the recall per patient is 100%, ensuring that a sick person will be classified as positive. As mentioned earlier, this application is intended to assist in medical diagnosis by providing an automatic pre-classification of patients, highlighting those who should receive special attention, but the final decision will always be made by the expert radiologist.

The *Precision* with datasets containing the “real world” proportion of sick/healthy breasts is very low, especially compared to results obtained with balanced datasets. This is a normal phenomenon in neural network training (real world accuracy is less in the majority class that was decimated for training). To solve this, it would be necessary to train with bigger datasets, especially with more samples of sick breasts.

The final step is to analyze the entire dataset, so the classification of all mammograms is carried out. The results obtained are shown in Table 8.

**Table 8.** Results for 100% of whole dataset.

|  | **Per image** | **Per patient** |
| --- | --- | --- |
| *TN* | 39869 | 12688 |
| *TP* | 1080 | 488 |
| *FN* | 76 | 4 |
| *FP* | 13643 | 10642 |
| *Precision* | 7.34% | 4.38% |
| *Recall* | 97.30% | 99.19% |
| *F<sub>1</sub>-score* | 13.64% | 8.40% |
| *Accuracy* | 74.97% | 55.31% |

The final results are as expected, with the *Recall* remaining high and close to 99%. Out of the entire image dataset, the program only classified 4 patients as false negatives.

## 4. Conclusions and future lines of work

### 4.1. Conclusions

With the aim of diagnosing breast cancer accurately and reducing the use of more invasive techniques, this project carried out the study and application of a Deep Learning technique for classifying mammograms according to whether they have cancer or not. More specifically, the design of a CNN that extracts features from training images and makes predictions for validation images.

All the experiments mentioned in the paper were conducted to evaluate which network yields the best results. In these trials, several network parameters were tested, i.e., the number of filters, the number of convolutional and max-pooling layers, and training options such as the number of training epochs needed.

The appropriate resolution for achieving efficient training was also evaluated, concluding that very similar results are obtained with images of 512×512 and 256×256. Therefore, the latter was chosen due to faster processing.

Experiments were conducted to evaluate different image pre-processing types and the proposed oversampling methods. Oversampling is necessary due to the high imbalance of images between the healthy and sick classes.

In the case of image preparation prior to training, two different pre-processing types were carried out to homogenize the dataset as best as possible since the sizes, grayscale histogram, contrast of the images (and even the hospitals from which they were extracted) are different. Both consisted of similar processes, with the difference that in the second, the images were deformed so that the breast occupied the entire image, providing more valuable information. In general, the results obtained are similar with both types of pre-processing, although in the case of *Recall*, the highest values are achieved with pre-processing 1.

Two types of oversampling were also included, as mentioned earlier, to mitigate as much as possible the high imbalance between the healthy and sick classes. Both aim to increase the minority class by doubling or tripling the original images of the dataset. The proposed methods were *RandReplicateFiles*, obtained from Matlab Central, and *ForgeImage*, a function developed for the forensic and cybersecurity fields, adapted for this specific problem. Significant results are achieved with the first technique, reaching an *Accuracy* of 93.15%, *Precision* of 89.49%, *Recall* of 97.78%, and *F_1_-score* of 93.45%.

These good results, along with the motivation that advances in this type of Deep Learning technology can greatly assist in the early diagnosis of diseases and even support specialists in decision-making, led to the implementation of a Matlab program that can potentially be applied in the future for help in breast cancer diagnosis. This program achieved significant results, reaching a *Recall* of 100% (99% in most cases), meaning it is capable of classifying all sick patients as positive for cancer.

### 4.2. Future lines of Work

In light of the results obtained in this work, some improvements are suggested that could be successfully implemented in the proposed program.

In the developed model, it is necessary to combine the existing data with other datasets to naturally balance the dataset and achieve a larger number of data to work with, as one of the main issues addressed was the imbalance between classes.

At present, the overall program is not capable of correctly classifying all negative mammograms, due to the high disparity between the number of healthy and diseased mammograms. Consequently, the number of false positives is high, resulting in lower *Precision* and *F_1_-score* values compared to *Recall*, which is close to 100%.

Although the results obtained from training the network were very good, there are many possible configurations, so further experimentation could be done to optimize the number of convolutional layers.

Currently, the idea of trying different pre-processing and oversampling methods from the ones proposed in this work is also being considered.

In the future, it might be worth considering transferring this program to Python.

## Data Availability

All data produced in the present study are available upon reasonable request to the authors.

https://www.kaggle.com/competitions/rsna-breast-cancer-detection/overview

## Notes

### Competing Interest Statement

The authors have declared no competing interest.

### Funding Statement

This study did not receive any funding

### Author Declarations

The study used ONLY openly available human data that were originally located at: https://www.kaggle.com/competitions/rsna-breast-cancer-detection/overview

